# Alcohol septal ablation in hypertrophic obstructive cardiomyopathy: do sex, mitral valve leaflet length and septal thickness affect the outcome?

**DOI:** 10.1101/2024.01.17.24301419

**Authors:** Mesud Mustafic, Rebecka Jandér, David Marlevi, Anette Rickenlund, Andreas Rück, Nawzad Saleh, Sam Abdi, Maria J Eriksson, Anna Damlin

## Abstract

**Purpose:** This retrospective cohort study aimed to assess whether basal septal wall thickness (BSWT), anterior (AML) and posterior (PML) mitral leaflet length, or sex were associated with remaining left ventricular outflow tract obstruction (LVOTO) in patients with hypertrophic obstructive cardiomyopathy (HOCM) undergoing alcohol septal ablation (ASA).

**Methods:** 154 patients that underwent ASA at the Karolinska University Hospital in Stockholm, Sweden, between 2009 and 2021, were retrospectively included. Anatomical and hemodynamic parameters were collected from invasive catheterization before and during ASA, and from echocardiography (ECHO) examinations before, during, and at one-year follow-up after ASA. Linear and logistic regression models were used to assess the association between sex, BSWT, AML, PML, and outcome defined as remaining LVOTO (≥30 mmHg) after ASA.

**Results:** The median follow-up was 364 days (IQR 334-385 days). BSWT ≥23 mm (n=13, 12%) was associated with remaining LVOTO at follow-up (*p=0.004*). Elongated MVLL (either AML or PML) was present in 125 (90%) of the patients. Elongated AML (>24 mm) was present in 67 (44%) of the patients, although AML length was not associated with remaining LVOTO at follow-up. Elongated PML (>14 mm) was present in 114 (74%) and not associated with remaining LVOTO at follow-up. No significant sex differences were observed regarding remaining LVOTO.

**Conclusion:** ECHO measurement of BSWT can be used effectively for patient selection for successful ASA and for identification of patients with a risk of incomplete resolution of LVOTO after ASA.

## Introduction

Hypertrophic obstructive cardiomyopathy (HOCM) is the most severe form of hypertrophic cardiomyopathy (HCM) where basal septal hypertrophy contributes to left ventricular outflow tract obstruction (LVOTO) and the patients also have increased risk of arrhythmias [1]. In addition to the hypertrophy of the septum, systolic anterior motion and abnormalities of the mitral valve leaflets are other known pathophysiological mechanisms behind LVOTO in patients with HOCM [2, 3]. Pharmacological treatment in HOCM aims to reduce the symptoms associated with LVOTO [1]. In instances where pharmacological treatment cannot mitigate severe symptoms and disease progress, alcohol septal ablation (ASA) is one of the recommended interventions for treatment of HOCM and LVOTO. ASA is a percutaneous procedure where a regional percutaneous alcohol injection is performed to induce a focal myocardial infarction in a selected part of the basal left ventricular septal wall, rendering a reduced septal wall thickness and LVOTO [4]. Compared to alternative surgical procedures such as myectomy, ASA is associated with shorter recovery time, faster intervention time, and fewer risks associated with cardiac surgery [5]. However, the proportion of patients that do not respond to ASA leading to further re-interventions is higher among patients undergoing ASA (5-25%) compared with surgical myectomy (<1%) [6, 7]. The reason for the higher re-intervention rate is not entirely understood.

Additional to the hypertrophic septum, patients with HCM may have elongated mitral valve leaflets in comparison to healthy controls, inducing further risk of developing LVOTO [3]. However, the impact of sex, pre-procedural basal septal wall thickness (BSWT) and mitral valve leaflet length (MVLL) on success rate in patients with HOCM undergoing ASA has been less studied. Therefore, this study aims to investigate if sex, BSWT and MVLL, are associated with remaining LVOTO in patients with HOCM, undergoing ASA.

## Methods

### Study design and population

This retrospective cohort study of all patients with HOCM that underwent ASA on clinical indication between June 2, 2009, and July 1, 2021, was conducted at the Karolinska University Hospital in Stockholm, Sweden, in total 158 patients. All patients were accepted for ASA procedure by a multi-disciplinary HCM team, consisting of clinical- and interventional cardiologists, clinical physiologists, and thoracic surgeons, based on clinical symptoms and significant LVOT-gradient documented by rest or exercise transthoracic Doppler-echocardiography (ECHO) despite optimal medical therapy. All patients had undergone a pre-procedural ECHO performed at maximum one week prior to ASA. One to three follow up visits with ECHO were performed during the first year after ablation, with additional follow-up if clinically motivated. In this study, if the one-year follow-up ECHO was not available, the latest study from the interval of 3-24 months post-intervention was included. Patients that were planned for an ASA but were not able to fulfill the procedure due to lack of suitable septal arterial branch were excluded (n=4) rendering a total of n=154 patients for the final analysis.

### Data collection

#### Patient data

Patient data as well as ASA specific data were collected from patient records, and from imaging systems: left-heart catheterization (CATH) (PACS), and ECHO data (Viewpoint). The data for each patient were collected from the three different time points; from the pre-procedural ECHO performed at maximum one week prior to ASA; CATH and ECHO during the ASA, and a follow up ECHO one year after ablation. Any values missing from the patients’ files were registered as missing values.

#### ECHO

All ECHO examinations were performed by personnel highly experienced in cardiac imaging using advanced ultrasound machines for cardiac imaging. The image acquisition was performed according to standard protocol including additional images focusing on HCM specific morphological and Doppler findings (as described below) to enable comprehensive HCM evaluation [8].

#### Pre-procedural ECHO

The data collected from the pre-ECHO was: age, sex, heart rate (beats per minute, BPM), weight, length, body surface area (BSA), LV end-diastolic and end-systolic diameters, LV end-diastolic and end-systolic volume, left atrium (LA) end-systolic volume, LV BSWT (Fig. 1) and posterior wall thickness [9]. Further, the grade of mitral- and aortic regurgitation, presence of SAM and LVOT peak systolic velocity and gradients at rest and Valsalva, estimated from LVOT velocity using the simplified Bernoulli equation (ΔP=4V^2^) were collected. For MV specific assessment, the length of the MV leaflets was measured [8] from the preoperative ECHO images (Fig. 1). The morphological measurements of the anterior mitral leaflet (AML) and posterior mitral leaflet (PML) length were made in the apical three-chamber view (A3C) at end-diastole (n=142, 92%). Among patients with poor visualization in A3C, the measurements were done in parasternal long axis view (PLAX) (n=9, 6%). Two ECHO examinations had too poor visualization to allow the MV specific measurements, these MV specific data were registered as missing (n=2, 1%). Data from one patient where the ECHO images were inaccessible were registered as missing (n=1, 1%).

**Fig. 1.**
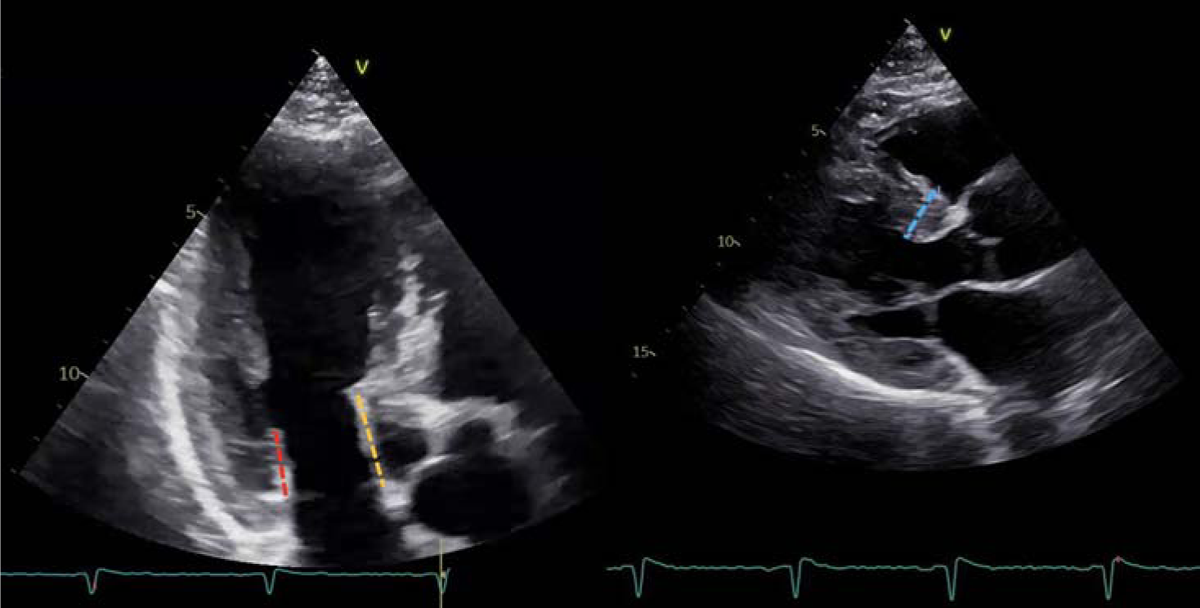
Measures of mitral valve leaflet length and basal septal wall thickness in pre-procedural ECHO images. A. Mitral valve leaflet length, red line: posterior mitral leaflet length, yellow line: anterior mitral leaflet length. B. Basal septal wall thickness, blue line: basal septal wall thickness.

#### Periprocedural invasive pressure measurements

Invasive intracardiac pressure measurements were performed using the standard end-hole fluid-filled catheters and recorded and stored using Exper Cardio System (Philips Healthcare). Data from pressure measurement were collected continuously throughout the ASA. The pressure gradients were measured invasively and simultaneously during CATH with one catheter placed in the LV and one in the ascending aorta. The following data was collected: heart rhythm, heart rate, LV peak systolic, early- and late diastolic pressures, peak systolic and diastolic pressures in the ascending aorta, both at rest and at Valsalva maneuver (Valsalva) and/or during induced premature ventricular contractions (PVC). Peak-to-peak systolic pressure gradient across LVOT was calculated as the difference between LV peak systolic pressure and aortic peak systolic pressure. Resting and provoked LVOT peak-to-peak gradients were calculated prior to, and 10 minutes after alcohol administration at the end of the ASA procedure. The amount of injected alcohol was registered.

#### Follow-up

The following data was collected from the one-year follow-up: average days until follow-up, pacemaker or ICD implantation, number of re-interventions (ASA or myectomy). ECHO data collected from the one-year follow-up: prevalence of SAM, BSWT, LV end-diastolic diameter (LVEDd), maximal LVOT velocity and gradients at rest and Valsalva, estimated from maximal LVOT velocity using the simplified Bernoulli equation (ΔP=4V ^2^). Remaining LVOTO was defined as a peak instantaneous gradient across LV outflow of at least 30 mmHg, either at rest or on provocation derived from the ECHO examination at the one-year follow-up, or from the most recent ECHO examination before re-intervention for the patients that underwent re-intervention.

#### Statistical analysis

To verify if continuous data were normally distributed, Shapiro-Wilk test was used. Normally distributed data were presented with mean ± standard deviation (SD). Non-normally distributed data were presented with median and interquartile range (IQR). Binary variables were reported as absolute numbers and percentages. To evaluate the correlation between sex, the AML length, the PML length, the BSWT, and the amount of alcohol injected, and outcome (remaining LVOTO) after ASA, logistic and linear regression models were executed. The linear regression models were adjusted for sex and age. To assess a cutoff value if any BSWT was associated with better outcome at follow-up, a number of cutoffs ranging from the different values in the IQR of BSWT were tried using logistic regression. P-values <0.05 were considered significant. Inter- and intra-observer variability of measures of BSWT, length of AML and PML, was calculated using intra-class correlation (ICC). Data analysis was conducted using STATA software (version 17.0 Stata Corp., College Station, Texas, USA).

## Results

### Baseline characteristics and pre-procedural ECHO

The baseline characteristics from the pre-procedural ECHO are presented in Table 1. The study population consisted of 77 men and 77 women. The pre-procedural ECHO was done at maximum 7 days before ASA, although 95% of the patients underwent the pre-procedural ECHO at the same day, or one day prior to the ASA. The pre-procedural ECHOs showed that 147 (95%) patients had SAM (with or without septal contact) at rest, however the patients without SAM at rest developed LVOTO with SAM at Valsalva maneuver. The median BSWT prior to the procedure was 19 mm (IQR 17-21 mm). The mean lengths of the AML and PML were 24.2 ± 3.2 mm, and 16.3 ± 2.8 mm, respectively. The mean amount of alcohol injected during ASA was 1.6 ± 0.3 milliliters (IQR 1.5-1.8 ml) and the mean amount per millimeter BSWT was 0.85 ± 0.2 mL/mm.

**Table 1.**
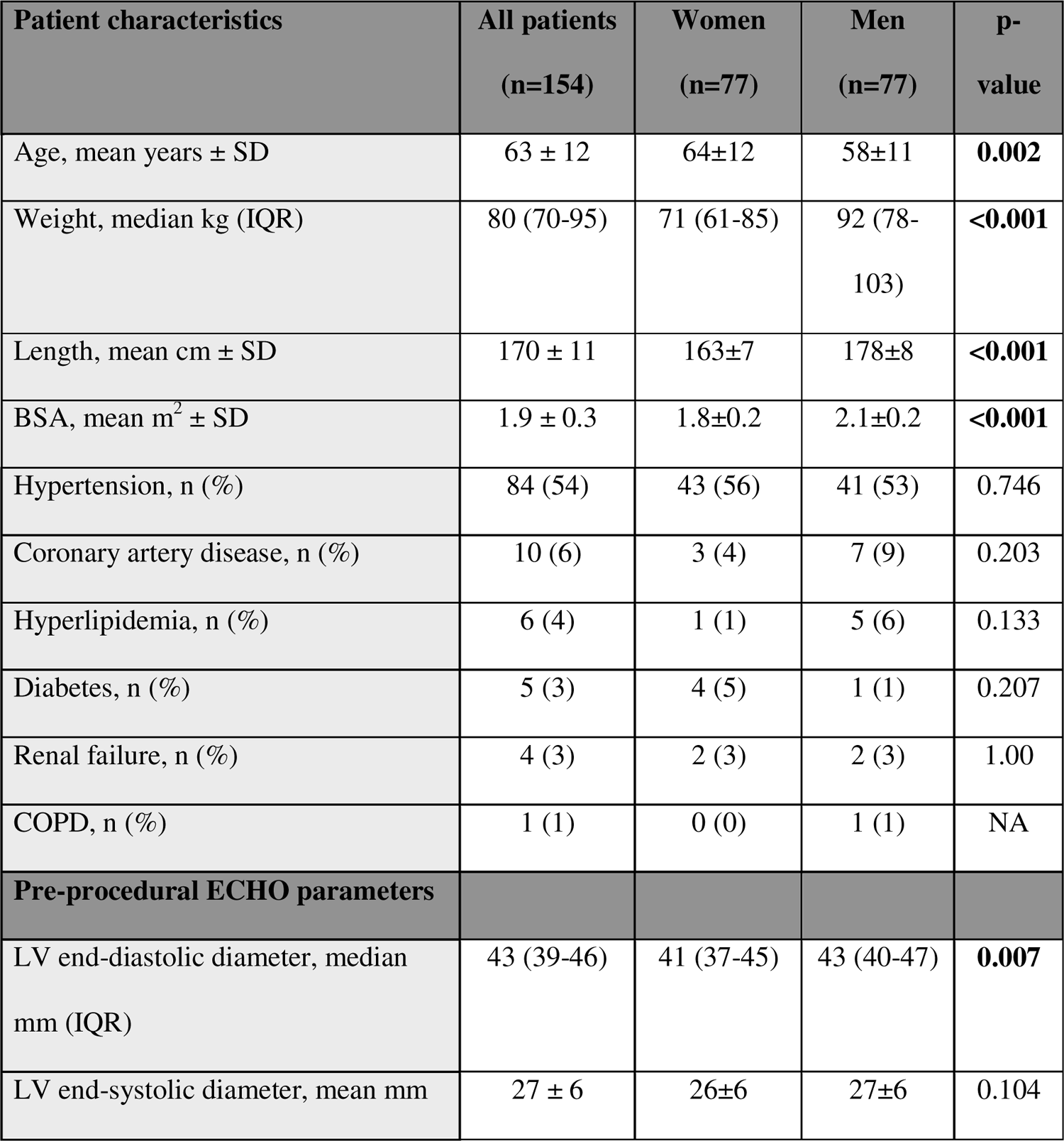
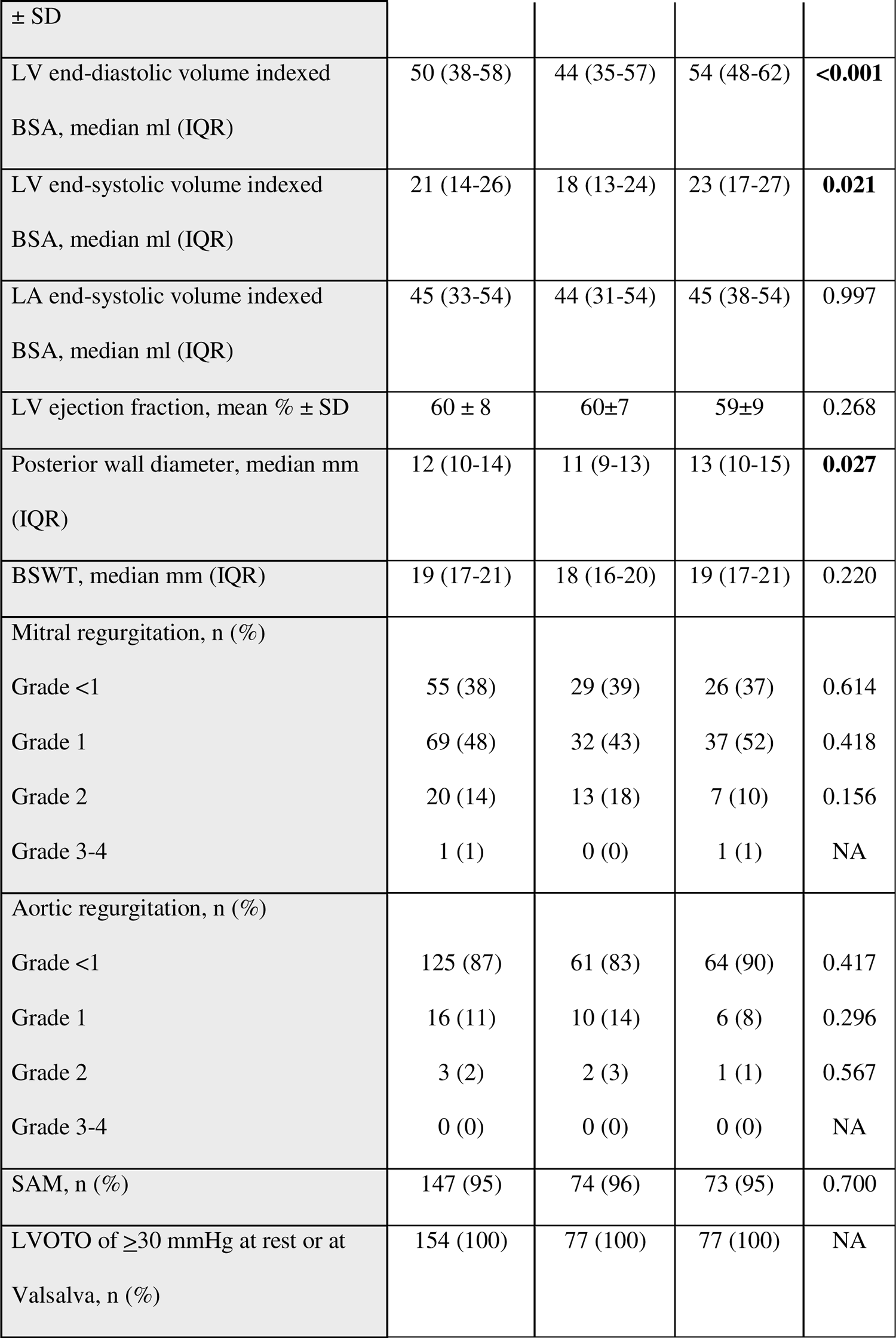
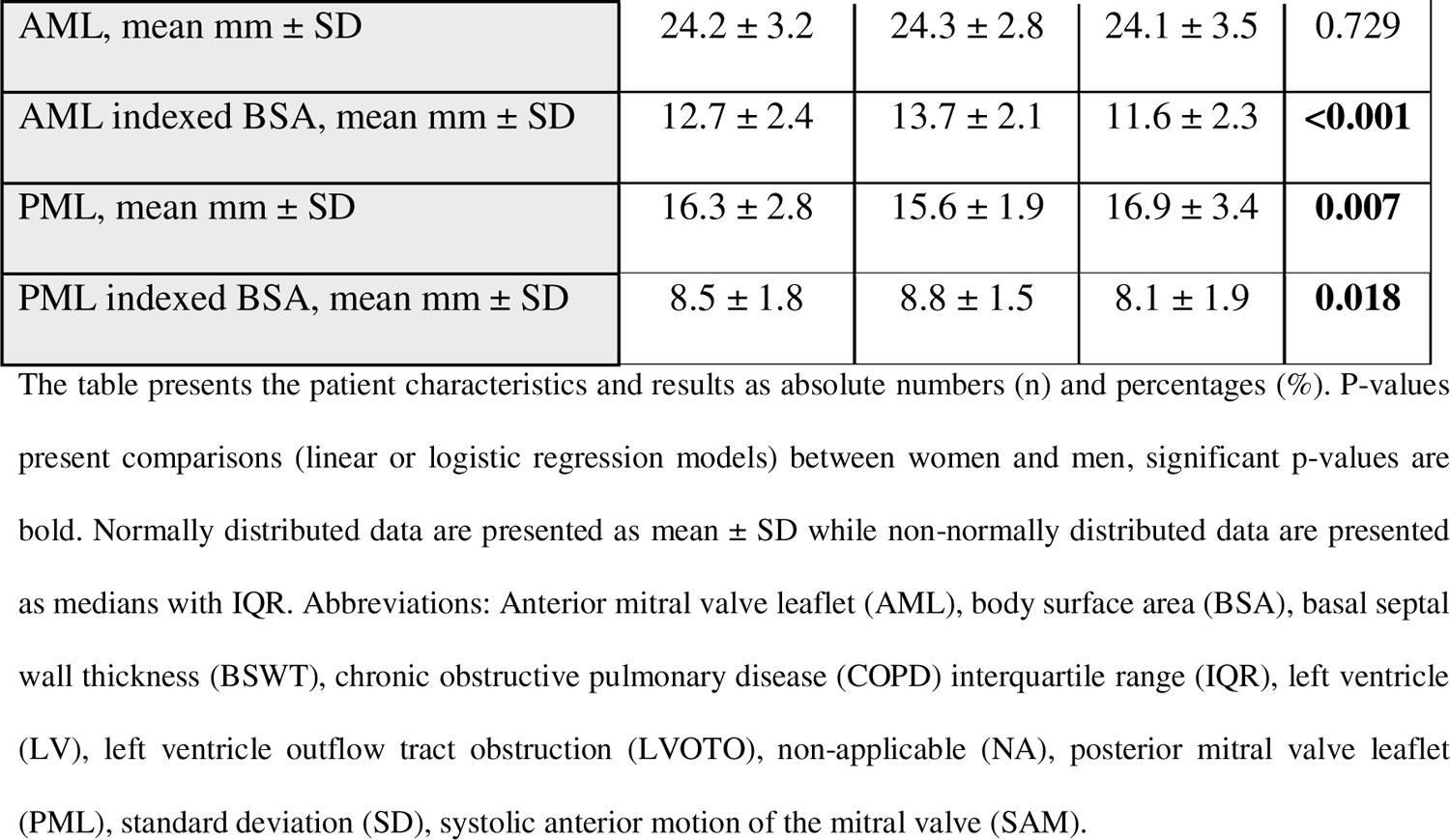
Patient characteristics and pre-procedural ECHO variables among patients with hypertrophic obstructive cardiomyopathy undergoing alcohol septal ablation.

### Intra-procedural invasive measurements and acute hemodynamic effects of ASA

The systolic pressures and peak-to-peak systolic pressure gradients pre- and post-ASA at rest and at Valsalva/PVC are presented in Figure 2. There was no significant change in the systolic aortic pressure pre- and post-ASA, however there was a significant decrease in the LV peak systolic pressure and LVOT peak-to-peak pressure gradient post-ASA compared to pre-ASA, at both rest and Valsalva.

**Fig. 2.**
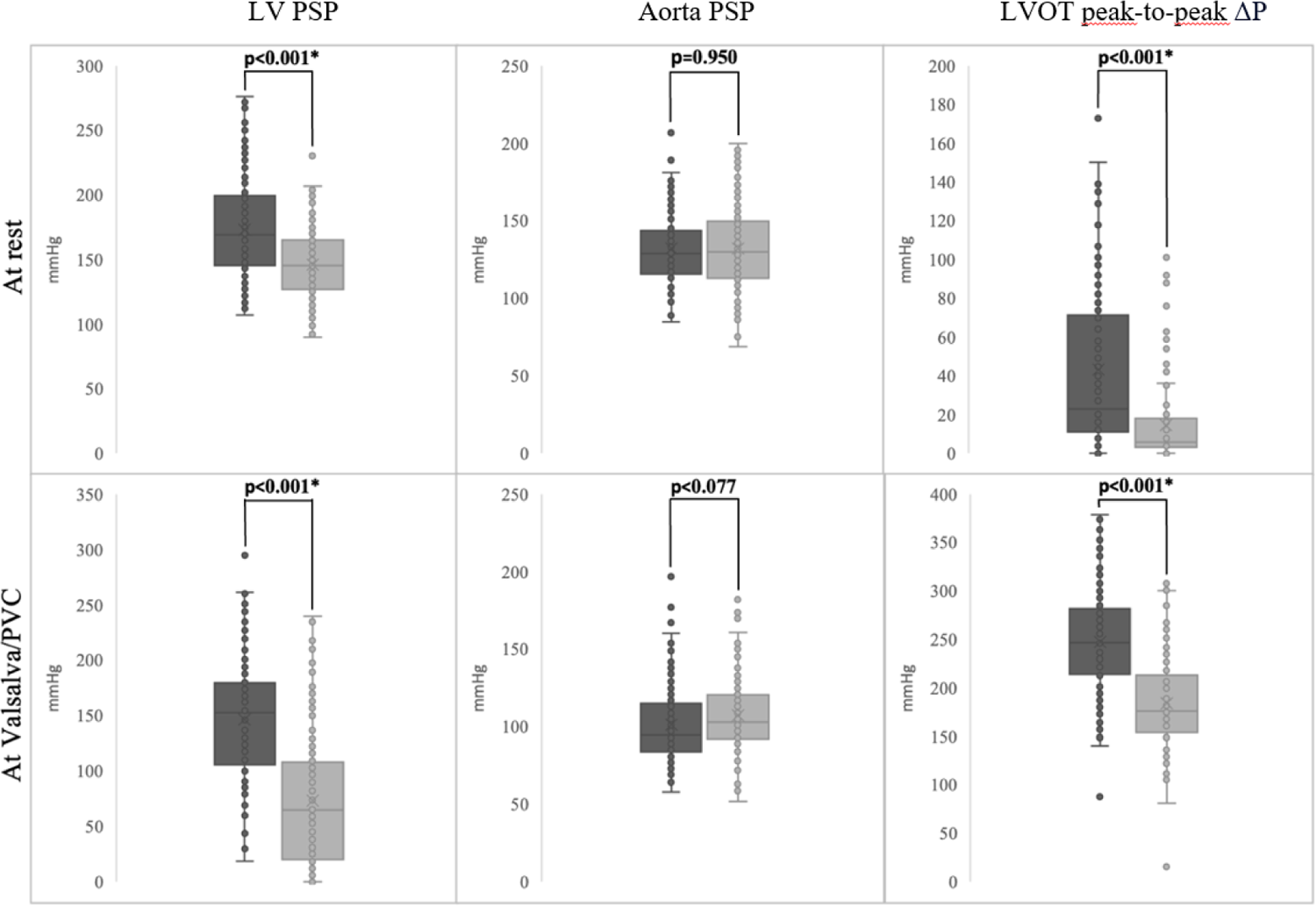
Comparison between invasive systolic left ventricular pressure gradients before (dark grey) and after (light grey) alcohol septal ablation at rest and during Valsalva/PVC. Values are expressed as median and interquartile ranges. *Statistically significant p values. Abbreviations: LV – Left ventricle, LVOT – Left ventricular outflow tract, PSP – peak systolic pressure, PVC – Induced premature ventricular contraction.

### Clinical outcomes and follow-up echocardiography

The follow-up ECHO was performed median 364 days post-procedure (IQR 334-385 days). During this follow-up period, 13 (8%) patients underwent a new ASA, and 1 (1%) patient required post-procedural myectomy. The patients that underwent re-intervention had the most recent ECHO examination before re-intervention registered as their follow-up ECHO. Between the ASA and the follow-up, 34 patients had got either a permanent PM (n=29, 19%) or ICD (n=5, 3%) implantation. The results from the follow-up ECHO examinations showed that 128 (86%) of the patients no longer had LVOTO after ASA. The amount of alcohol injected during the procedure was not associated with remaining LVOTO at follow-up (p=0.865).

There was a significant reduction in septal thickness from the pre-procedural ECHO and the follow-up ECHO (from pre-procedural 19 mm (IQR 17-21 mm) to 16mm (IQR 13-18 mm) at follow-up (p<0.001). Of all patients, 19 (12%) had BSWT ≥23 mm, which was significantly associated with remaining LVOTO at follow-up (7 (37%) of the patients with BSWT ≥23 mm had remaining LVOTO at follow-up, compared to 11% of the patients with BSWT <23 mm, p=0.004, Table 2). Elongated MVLL (either AML or PML) was present in 125 (90%) of the patients. Elongated AML (>24 mm) was present in 67 (44%) of the patients, although AML length was not associated with remaining LVOTO at follow-up. Elongated PML (>14 mm) was present in 114 (74%) of the patients but neither PML was associated with remaining LVOTO at follow-up (Table 2).

**Table 2.**
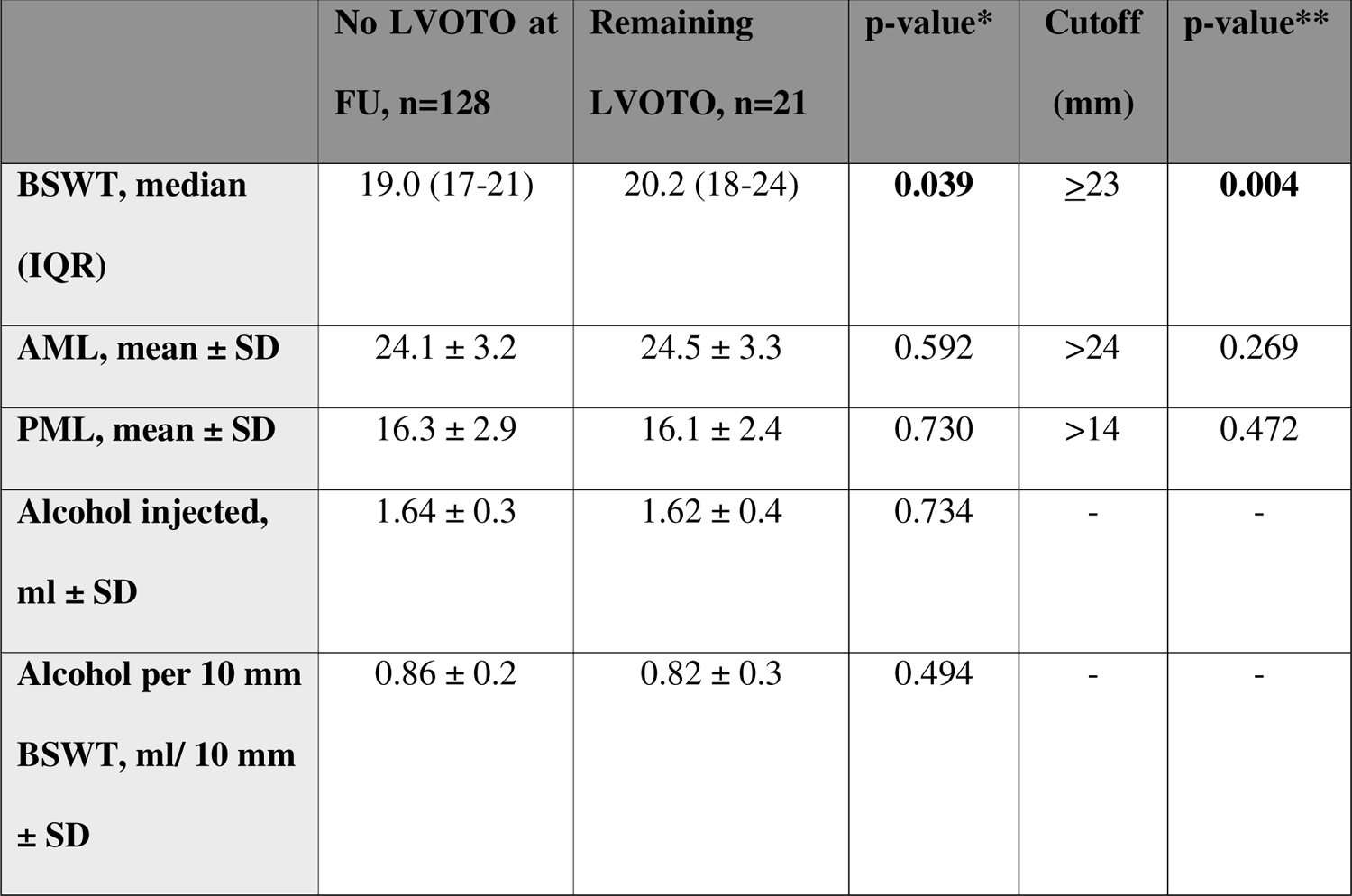
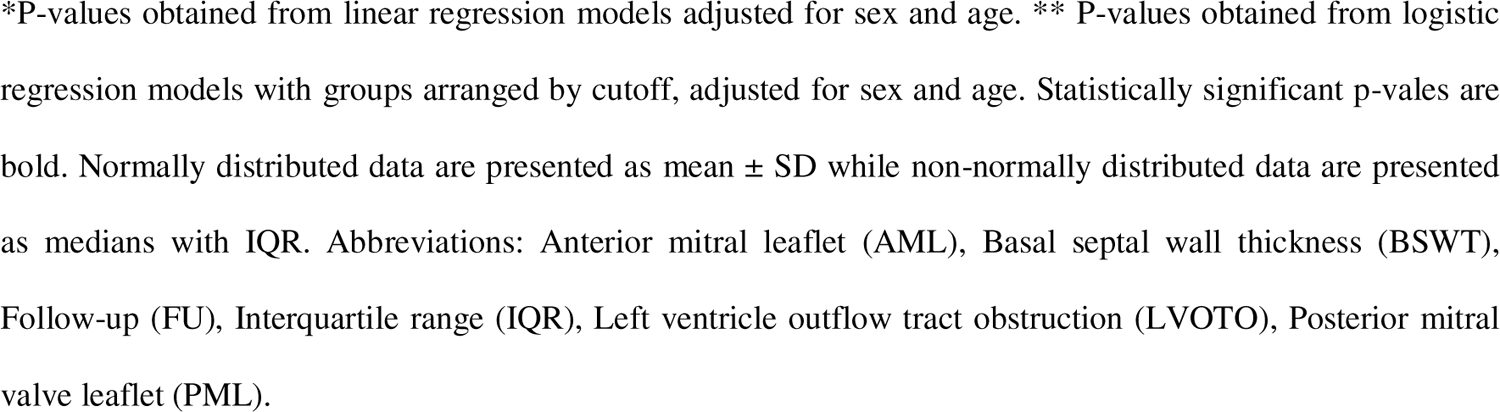
Basal septal wall thickness, anterior and posterior mitral leaflet length, and amount alcohol in relation to remaining left ventricular outflow tract obstruction and re-ablation after alcohol septal ablation.

|  | No LVOTO at<br>FU, n=128 | Remaining<br>LVOTO, n=21 | p-value* | Cutoff<br>(mm) | p-value** |
| --- | --- | --- | --- | --- | --- |
| **BSWT, median**<br>**(IQR)** | 19.0 (17-21) | 20.2 (18-24) | **0.039** | $\geq 23$ | **0.004** |
| **AML, mean ± SD** | 24.1 ± 3.2 | 24.5 ± 3.3 | 0.592 | >24 | 0.269 |
| **PML, mean ± SD** | 16.3 ± 2.9 | 16.1 ± 2.4 | 0.730 | >14 | 0.472 |
| **Alcohol injected,**<br>**ml ± SD** | 1.64 ± 0.3 | 1.62 ± 0.4 | 0.734 | - | - |
| **Alcohol per 10 mm**<br>**BSWT, ml/ 10 mm**<br>**± SD** | 0.86 ± 0.2 | 0.82 ± 0.3 | 0.494 | - | - |

\*P-values obtained from linear regression models adjusted for sex and age. \*\* P-values obtained from logistic regression models with groups arranged by cutoff, adjusted for sex and age. Statistically significant p-values are bold. Normally distributed data are presented as mean ± SD while non-normally distributed data are presented as medians with IQR. Abbreviations: Anterior mitral leaflet (AML), Basal septal wall thickness (BSWT), Follow-up (FU), Interquartile range (IQR), Left ventricle outflow tract obstruction (LVOTO), Posterior mitral valve leaflet (PML).

### Sex differences

There were no significant differences in outcomes between men and women neither when comparing CATH- or ECHO parameters (Table 3). There was no difference in remaining LVOTO between women and men, neither at post procedural CATH nor at follow-up (Table 3). Men had significantly longer PML compared with women, although both AML and PML length indexed for BSA was longer among women (Table 1).

**Table 3.** Sex differences at one-year follow-up after alcohol septal ablation in patients with hypertrophic obstructive cardiomyopathy.

| **Clinical Outcomes** | **All patients** | **Men (n=77)** | **Women (n=77)** | **P values** |
| --- | --- | --- | --- | --- |
| Myectomy, n (%) | 1 (1) | 0 (0) | 1 (1) |  |
| Re-ablation, n (%) | 13 (9) | 5 (7) | 8 (11) | 0.375 |
| Pacemaker, n (%) | 29 (20) | 14 (18) | 15 (20) | 0.806 |
| ICD, n (%) | 5 (3) | 2 (3) | 3 (4) | 0.641 |
| **Post procedural CATH outcomes** |  |  |  |  |
| Remaining LVOTO at rest, n (%) | 29 (19) | 11 (14) | 18 (23) | 0.153 |
| At least 50% reduction of acute LVOT gradient at rest, n (%) | 68 (44) | 30 (40) | 38 (51) | 0.165 |
| Remaining LVOTO at Valsalva/PVC, n (%) | 111 (72) | 56 (73) | 55 (71) | 0.857 |
| At least 50% reduction of acute LVOT gradient at Valsalva/PVC, n (%) | 83 (54) | 40 (56) | 43 (60) | 0.682 |
| **Follow-up ECHO outcomes** |  |  |  |  |
| Remaining LVOTO, n (%) | 21 (14) | 8 (10) | 13 (17) | 0.230 |
| At least 50% reduction of LVOT gradient at rest, n (%) | 72 (47) | 32 (43) | 40 (54) | 0.165 |
| At least 50% reduction of LVOT gradient at Valsalva, n (%) | 82 (53) | 40 (54) | 42 (58) | 0.602 |
| Base septal thickness reduction, mm, mean±SD | -3.4 ± 4.1 | -3.3 ± 3.9 | -3.5 ± 4.1 | 0.700 |

Values are expressed as absolute numbers (n) and percentages (%) or mean±standard deviation for normally distributed variables. P-values were obtained from logistic regression models. The P value was left out for myectomy as no men underwent myectomy. Abbreviations: alcohol septal ablation (ASA), echocardiography (ECHO), Implantable cardioverter defibrillator (ICD), invasive intracardiac pressure examination (CATH), Left ventricle outflow tract (LVOT), Induced premature ventricular contraction (PVC), standard deviation (SD).

### Peri- and post-procedural complications

In total, 30 (19%) of the patients got either AV block II or III between the ASA and the one-year follow-up. AV block II or III were more common among women (n=20, 26%) compared with men (n=10, 13%, p=0.045). Four of the patients (3%) got atrial fibrillation after ASA, one patient (1%) had post-procedural tamponade, and one (1%) had cardiac arrest. One patient deceased within 30 days after ASA, and within one year after the ASA additionally three patients deceased. There were no sex differences in mortality (p=0.225).

### Intra- and interobserver analysis

Intra- and interobserver agreements for measurements of BSWT and MVLL were analyzed and are presented in Table 4.

**Table 4.** Intra- and interobserver agreement.

|  | Interobserver | Intraobserver |
| --- | --- | --- |
|  | ICC | ICC |
| BSWT | 0.92, p=0.001 | 0.89, p=0.001 |
| AML | 0.65, p=0.035 | 0.83, p=0.004 |
| PML | 0.89, p=0.001 | 0.93, p<0.001 |

Intraclass correlation coefficients (ICCs) were obtained using two-way mixed-effects models as a measure of agreement. Abbreviations: Anterior mitral valve leaflet (AML), basal septal wall thickness (BSWT), intraclass correlation coefficients (ICC), posterior mitral valve leaflet (PML).

## Discussion

This single-center retrospective cohort study aimed to assess if there were any associations between sex, BSWT, AML length, PML length, and outcomes after ASA-treatment for HOCM. In general, patients with BSWT ≥23 mm had a significantly higher rate of remaining LVOTO at follow-up. Most patients had elongated MVLL, however AML and PML length were not associated with remaining LVOTO at follow-up. Further, the amount alcohol injected during ASA was not associated with remaining LVOTO at follow-up. These results suggest that BSWT could be a possible predictive value for LVOTO reduction after ASA, however there is a need for further studies of potential anatomical predictors of HOCM-patients, to enable optimized treatment based on individual conditions.

At the one-year follow-up, 14% of the patients had remaining LVOTO, which is adherent to a recent review of ASA showing 10-20% of patients have a residual gradient > 30 mmHg after ASA [10]. Supportive to this study, showing that BSWT ≥23 mm had a significantly higher rate of remaining LVOTO at follow-up, it has previously been shown that BSWT is associated with LVOTO in patients with HOCM [11]. Further supportive, a study of 531 HOCM-patients with follow-up time 0.6±0.6 years after ASA, showed that patients with thicker septum had a higher residual LVOTO on follow-up compared to patients with a thinner septum [12]. That study had 3 cutoffs: <20mm, 20-24mm, ≥25mm, where the ≥25mm group showed a trend of higher residual gradients on follow-up although of no significance compared to the other groups [12]. Additionally, a study by Lu *et al*. with 102 HOCM patients found that a septum thinner than 24.3 mm had a higher rate of LVOT gradient reduction defined as a 50% reduction compared to the gradient before ASA, and patients with a septal thickness of ≥24.3 mm had more non-responders [13]. The authors suggested having 24.3 mm as a cutoff when considering treating patients with ASA since a significant number of patients with a septum thicker than that were non-responders and had worse outcome in form of LVOT gradient reduction [13]. Their study had a different definition of LVOT reduction, with a 50% reduction in LVOT gradient between pre procedural examination and follow-up. In summary, BSWT seems to be a promising predictive value for LVOTO reduction after ASA.

In this study, the amount of alcohol injected during the procedure was not associated with remaining LVOTO at follow-up. The mean amount of alcohol injected during the ASA in this study (1.6 ± 0.3 mL) is lower than in previous studies reporting mean alcohol doses of 2.1-2.2 mL [14]. However, the BSWT this study was lower (mean 19 mm) than in the study for comparison (20.1-20.7 mm). Although the recommended dose of alcohol in ASA is 1 mL per 10 millimeters of BSWT [15], the amount of alcohol per 10 millimeters of BSWT is this study equaled 0.85 ± 0.2 mL/mm, which is lower than the recommended dose. This could possibly explain some of the cases of remaining LVOTO after ASA. At the one-year follow-up, one patient had undergone surgical myectomy and 13 (9%) patients had undergone re-ablation. Hence, 7 patients with remaining LVOTO after ASA did not undergo re-intervention during the first year after ASA, although some of these patients could have undergone re-intervention later than one year after ASA or they might have had a relief of symptoms as the LVOT gradient could have been reduced although > 30 mmHg.

Elongation of the AML and PML (as compared to normal reference range [11]) was observed in most patients in this study, in-line with previous studies of patients HOCM suggesting elongated MV leaflets as being part of the innate HCM phenotype [11,16–18]. Although the correlation between MVLL and outcome after ASA has not yet been shown. Lentz Carvalho *et al.* examined the effect of AML length in relation to outcomes after septal myectomy in 564 patients with HOCM and suggested that AML length ≥ 30 mm had no correlation with remaining LVOTO [3]; however, the study did not extent to assessment of possible sex differences. The finding that BSA-indexed valve leaflet lengths were significantly longer in women in this study, highlight a possible persistent functional difference that need to be further studied. Further, this study presents that women had a higher prevalence of post-operative complications (i.e. atrioventricular block) than men. This is supported by a study by Saravanabavanandan *et al.* shown that women have higher short-term all-cause mortality, incidence of atrioventricular block, permanent pacemaker implantation and hospital stay after ASA or myectomy, compared to men [19]. In both, this study and the study by Saravanabavanandan *et al.,* women were older at the time of the procedure.

### Strengths and Limitations

This study was performed as a retrospective cohort study examining patients diagnosed with HOCM undergoing ASA. A strength of the study was the amount of data collected and reported for all included patients, enabling an evaluation with few excluded data points. Further the study population had an equal gender distribution, and the number of patients (n=154) was satisfactory. The study specific variables i.e., AML and PML lengths were measured by two observers according to the previously described methodology and discussed in the team if needed. The measuring methodology was in correspondence with previous studies (7, 9, 17). A potential limitation could be that the examinations were performed by different examiners which may affect the repeatability of the results. Additionally, in this study two patients, equal to 1%, were excluded from the MV specific measurements due to inadequate ECHO quality. In the pre- and post-procedural examinations, a total of 14 patients (9%) were measured in PLAX view instead of A3C view, due to poor visualization which could have only a limited impact on the results. The ICC values indicate good reliability for BSWT and PML length measurements, however on a more moderate level for AML length.

The length of AML can be sometimes difficult to measure in HOCM, often due to calcification in the PML base. We have also shown that patients with a basal interventricular septum equal and thicker than 23 mm had a lesser reduction of LVOTO after ASA, and therefore might need a longer follow-up for re-evaluation.

## Conclusion

This study suggests that pre-procedural septal thickness ≥23 mm was associated with remaining LVOTO at follow-up one year after ASA. Length of AML and PML, and sex do not seem to contribute significantly to remaining LVOTO. Most patients with HOCM had elongated MVLL, which supports the notion of elongated MV leaflets in patients with HOCM. In summary, these findings suggest that BSWT measured with ECHO can be used effectively for patient selection for successful ASA. A severe basal septal hypertrophy in HOCM may be associated with a poor outcome after ASA. There is a need for further studies of potential sex differences and detectable differences in the anatomy of HOCM-patients, to enable optimized treatment based on individual conditions.

## Statements & Declarations

### Funding

D.M. would like to acknowledge funding from the Swedish Heart-Lung foundation (project number 20230080), and the European Union (ERC, MultiPRESS, 101075494). Views and opinions expressed are those of the authors and do not reflect those of the European Union or the European Research Council Executive Agency. A.D. would like to acknowledge funding from the Swedish Heart-Lung foundation (project number 20230742).

### Competing interests

A.D has received minor speaker honorarium from Edwards Lifesciences.

### Author contributions

All authors contributed to the study conception and design. Material preparation was performed by Anna Damlin, Anette Rickenlund, Maria J Eriksson, Andreas Rück and Nawzad Saleh. Data collection and analysis were performed by Mesud Mustafic, Rebecka Jandér, Sam Abdi, David Marlevi, and Anna Damlin. The first draft of the manuscript was written by Mesud Mustafic and Rebecka Jandér under supervision of Anna Damlin, and all authors commented on previous versions of the manuscript. All authors read and approved the final manuscript.

### Ethics approval

This study was performed in line with the principles of the Declaration of Helsinki. Approval was granted by the Swedish Ethical Review Authority with the registration number: 2022-01472-01.

### Consent to participate

This study does not present any identifying details. All information was anonymized, and the study did not imply any risks for the patients since it did not affect the patient or cause any changes in their treatments or care.

## Data Availability

All data produced in the present study are available upon reasonable request to the authors.

## Notes

### Competing Interest Statement

The authors have declared no competing interest.

### Funding Statement

Funding from the Swedish Heart-Lung foundation (project number 20230080 and project number 20230742), and the European Union (ERC, MultiPRESS, 101075494).

### Author Declarations

Approval was granted by the Swedish Ethical Review Authority with the registration number: 2022-01472-01.

